# Psychological Impact of COVID-19 on Pakistani University Students and How They Are Coping

**DOI:** 10.1101/2020.05.21.20108647

**Authors:** Muhammad Salman, Noman Asif, Zia Ul Mustafa, Tahir Mehmood Khan, Naureen Shehzadi, Khalid Hussain, Humera Tahir, Muhammad Husnnain Raza, Muhammad Tanveer Khan

**Affiliations:** Faculty of Pharmacy, The University of Lahore, 1-Km Defense road, Lahore, Pakistan; Punjab University College of Pharmacy, University of the Punjab, Lahore, Pakistan; Gulab Devi Educational Complex, Lahore, Pakistan; District Headquarter Hospital, Pakpattan, Pakistan; Institute of Pharmaceutical Science, University of Veterinary and Animal Sciences, Lahore, Pakistan; Ruth Pfau College of Nutrition Sciences, Lahore Medical and Dental College, Lahore, Pakistan; Faisalabad Institute of Cardiology, Faisalabad, Pakistan.

**Keywords:** Anxiety, COVID-19, Coping, Depression, Pakistan, SARS-CoV2, University students

## Abstract

**Background:** COVID-19 is spreading quickly, causing great deal of fear and unrest in the public. We aimed to assess the psychological impact of COVID-19 on university students and their coping strategies.

**Methods:** This web-based, cross-sectional study was conducted among students of four Pakistani higher education institutions. Google forms were used to disseminate the online questionnaire to assess anxiety (GAD-7), depression (PHQ-9), sources of distress (14-items) and the coping strategies (Brief-COPE).

**Results:** A total of 1134 responses (age 21.7 ± 3.5 years) were included. The frequency of students having moderate-severe anxiety and depression (score ≥ 10) were ≈ 34% and 45%, respectively. The respondents’ aged ≥ 31 years had significantly lower depression score than those below 30 years. Males had significantly less anxiety and depression scores than females. Additionally, those having a family member, friend or acquaintance infected with the disease had significantly higher anxiety score. The main sources of distress were the changes in daily life due to the ongoing pandemic. Regarding coping strategies, majority of respondents were found to have adopted religious/spiritual coping (6.45 ± 1.68) followed by acceptance (5.58 ± 1.65).

**Conclusions:** COVID-19 have significant adverse impact on students’ mental health. The most frequent coping strategy adopted by them are religious/spiritual coping, acceptance, self-distraction and active coping. It is suggested that mental health of students should not be neglected during epidemics.

## 1. Introduction

Historically, emergence and re-emergence of large-scale infectious diseases (IDs) have had civilization-altering consequences. In addition to physical problems, a variety of psychosocial problems also emerges mainly due to the lack of sufficient knowledge about these IDs. Initially, the fear, anxiety and hysteria in people are observed that lead to stigma – irrational response to the disease - in the society. Recent examples in this context include stigmatization of HIV/AIDS, SARS, H1N1 and Ebola. Such impulsive reactions reveal the enormous psychological distress consequential of emerging diseases particularly when it is unfamiliar, highly contagious and fatal such as ongoing COVID-19 pandemic.

In Pakistan, the first case of COVID-19 appeared on 26^th^ February 2020. The situation escalated quickly and in order to effectively contain COVID-19, on 23rd March 2020, a complete lockdown was imposed in the country. This complete lock down was converted into ‘smart lockdown’ on 9^th^ May, 2020. However, all the education institutions as well as big markets and all public places were directed to remain closed (Kaleem, 2020).

The continuous spread of the disease, conspiracy theories, myths and blame games, sensational media reporting of COVID-19, frustration and boredom, implementation of social lockdown with classmates, friends, and teachers, lack of personal space at home, and family financial loss due to lockdown are some of the main risk factors significantly influencing the mental health of university students. There have been reports on the psychological impact of the epidemic on the general public, healthcare workers and college students (Wang et al., 2020; Cao et al., 2020; Chew et al., 2020; Li et al., 2020). However, to our knowledge, no studies have assessed the psychological impact of COVID-19 on Pakistani university students. Therefore, the present study was conducted to underscore the psychologic impact of CVOID-19 on Pakistani university students and their coping strategies.

## 2. Methods

### 2.1. Study design, settings and subjects

A web-based, cross-sectional survey was conducted among Pakistani university students in the month of April and May 2020. All the students acquiring education at University of the Punjab, The University of Lahore, Gulab Devi Educational Complex and University of Veterinary and Animal Sciences were eligible for inclusion in this study. We excluded those who were not university students, who were already graduated and those unwilling to take part in the study.

### 2.2. Ethical consideration

Protocol of the present study was reviewed and approved by the Research Ethics Committee of the Department of Pharmacy Practice, Faculty of Pharmacy, The University of Lahore. An informed consent was obtained from every study participant.

### 2.3. Data collection tool

Google forms were used to disseminate the online questionnaire among the student aiming to assess anxiety, depression, sources of distress and the coping strategies during the ongoing COVID-19 pandemic. The content of the questionnaire were reviewed by an expert panel and, after suggested changes have been made, were approved for data collection. Additionally, the questionnaire was piloted among ten university students (age 20-30 years; 4 males and 6 females). All the participants reported ease of understanding all the items and response options. Data of these participants were not included in the final study. As shown in Figure 1, the final questionnaire had five sections. Section-I included an informed consent sheet. Those who consented filled out the subsequent sections.

**Figure 1:**
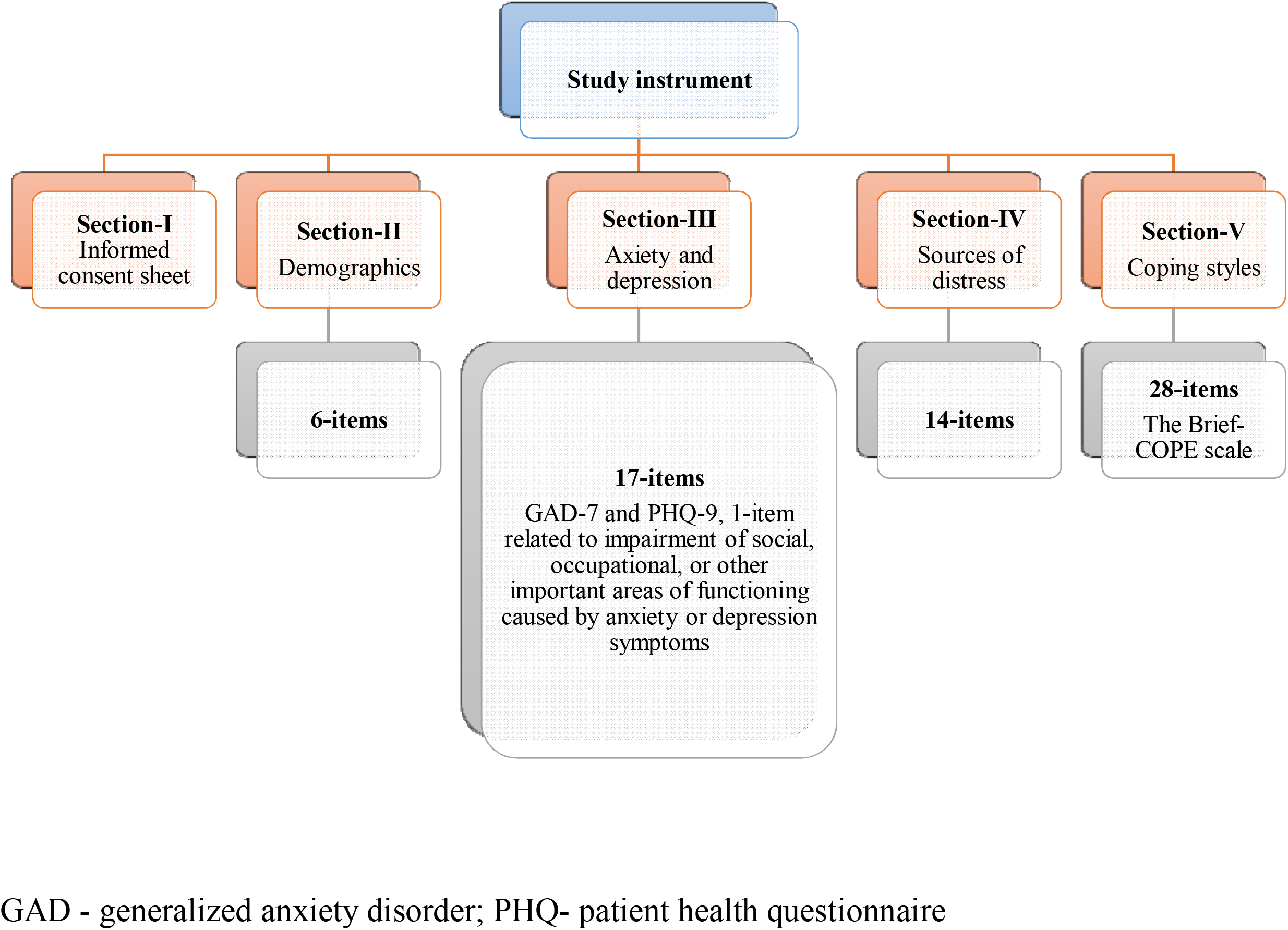
Sections of the study instrument GAD - generalized anxiety disorder; PHQ- patient health questionnaire

### 2.4. Outcome measures

In the present study, generalized anxiety scale (GAD-7) was used to assess anxiety (Spitzer et al., 2006). It contains seven items, each of which is scored 0 (not at all) to 3 (nearly every day), providing a 0 to 21 score. Scores of 5-9, 10-14, and ≥ 15 are taken as the cut-off points for mild, moderate and severe anxiety, respectively. Using a cut-off score of ≥ 10, the GAD-7 has asensitivity of 89% and a specificity of 82%. Moreover, it is also moderately good at screening three other common anxiety disorders: panic disorder (sensitivity 74% and specificity 81%), social anxiety disorder (sensitivity 72% and specificity 80%) and post-traumatic stress disorder (sensitivity 66% and specificity 81%).

Patient health questionnaire (PHQ-9) was used to screen depression in the study participants. PHQ-9 is one of the most commonly used instruments in practice as well as research. It contain 9-items each of which is scored 0 (not at all) to 3 (nearly every day), yielding a 0-27 score. PHQ-9 scores of ≤ 4, 5-9, 10-14, 15-19, and ≥ 20 are considered minimal, mild, moderate, moderately severe and severe depression, respectively (Kroenke et al., 2001).

The sources of distress from the ongoing COVID-19 epidemic were measured with a 14-item scale designed from a previous study reporting anxiety among university students during SARS outbreak (Wong et al., 2007). The respondents rated each item on a 3-point Likert-scale (−1 = disagree, 0 = neutral, and 1 = agree). The items were grouped under 4 scales namely fear of health of self, family and loved-ones (possible score −4 to 4), virus spread (possible score −3 to 3), effects on daily life (possible score −4 to 4), and discontent with measures taken by the government (possible score −3 to 3).

The Brief-COPE questionnaire was used to evaluate the coping strategies adopted by the study participants. It is a validated 28-items self-report questionnaire that measures effective and ineffective ways to cope with a stressful life event (Carver, 1997). Responses to each item are scored from one (I have not been doing this at all) to four (I have been doing this a lot). The Brief Cope explore the following 14 coping methods: self-distraction, active coping, denial, substance use, use of emotional support, use of instrumental support, behavioral disengagement, venting, positive reframing, planning, humor, acceptance, religion and self-blame. Possible scores ranged from 2 to 8 for each coping style; higher scores indicated a higher tendency to implement the corresponding coping style.

### 2.5 Statistical analysis

Responses stored in the web-based database (The Google Drive) of the principal investigator were transferred to Microsoft Excel sheet. After appropriate coding and data cleaning, the data were imported into the SPSS version 22 for the analysis. Continuous variables were expressed as mean and standard deviations (SD) whereas frequency and percentages were used to present categorical data. Independent t test and analysis of variance were used to determine significance for continuous data, where applicable, and Chi-Square test were performed for categorical data. A*p* value of less than 0.05 was taken for statistical significance.

## 3. Results

### 3.1. Characteristics of the study sample

A total of 1134 responses were included in this study. Upon analysis, it was revealed that majority (70.5%) of the respondents were female and native of Punjab province (93.4%), and 67.9% of the students were enrolled for the Doctor of Pharmacy program. Around 22% of the respondents disclosed of having a family member, relative, friend or acquaintance infected with the disease.

### 3.2. Anxiety and depression

The mean anxiety and depression score were 7.48 ± 5.65 and 9.42 ± 7.01, respectively. The frequency of students having moderate to severe anxiety (score ≥ 10) were ≈ 34%. Regarding the severity of depression, 30.5%, 24.5%, 21%, 13.6% and 10.4% students were found to have minimal-none, mild, moderate, moderately severe and severe depression, respectively. Between-demographic analysis of anxiety and depression scores are described in Table 1. The respondents age 31 years and above had significantly lower depression score in comparison to the age groups below 30 years. Similarly, male respondents were observed to have a significantly less anxiety and depression score than female respondents. In addition, those who reported of having family members, relative, friend or acquaintance who got contracted the disease had a significantly higher anxiety score *(p* < 0.001).

**Table 1:** Anxiety and depression assessment based on demographics of respondents

| Demographics | N (%) | Anxiety score | Depression score |
| --- | --- | --- | --- |
| **Age** [Mean 21.7 ± 3.5 years] |  |  |  |
| 18-20 years | 462 (40.7%) | 7.38 ± 5.57 | 9.82 ± 6.95 |
| > 20-25 years | 574 (50.6%) | 7.56 ± 5.72 | 9.35 ± 7.02 |
| > 25-30 years | 66 (5.8%) | 7.94 ± 5.72 | 8.64 ± 7.41 |
| ≥ 31 years | 32 (2.8%) | 6.53 ± 5.55 | 6.50 ± 6.17* |
| **Gender** |  |  |  |
| Male | 335 (29.5%) | 6.62 ± 5.70<sup>†</sup> | 8.73 ± 6.84* |
| Female | 799 (70.5%) | 7.84 ± 5.60 | 9.71 ± 7.06 |
| **Province** |  |  |  |
| Punjab | 1059 (93.4%) | 7.42 ± 5.62 | 9.35 ± 6.93 |
| Other provinces | 75 (6.6%) | 8.11 ± 5.88 | 10.17 ± 7.90 |
| **Education** |  |  |  |
| Medical | 43 (3.8%) | 7.19 ± 5.67 | 7.74 ± 6.59 |
| Pharmacy | 770 (67.9%) | 7.48 ± 5.74 | 9.52 ± 6.99 |
| Allied Health Sciences | 62 (5.5%) | 7.19 ± 5.37 | 8.32 ± 6.27 |
| Others | 259 (22.8%) | 7.60 ± 5.47 | 9.67 ± 7.27 |
| **Family member, relative or acquaintances got COVID-19** |  |  |  |
| Yes | 247(21.8%) | 8.89 ± 5.74<sup>†</sup> | 10.06 ± 6.91 |
| No | 887 (78.2%) | 7.09 ± 5.56 | 9.24 ± 7.03 |

\*P-value < 0.05; \*\*P-value < 0.01; <sup>†</sup>P-value < 0.001; One-way ANOVA was applied for three or more groups. To estimate the difference among two groups independent sample t-test was applied. P-value <0.05 was considered statistically significant.

### 3.3 Sources of distress

As shown in Table 2, the main sources of distress were the adverse effect of COVID-19 on daily life followed by the virus spread, and dissatisfaction with measures taken by the government to combat the disease. Regarding the fear of health of self and family members, majority (70.9%) of the respondents expressed fear of their family members and friends getting infected. However, around 41% were afraid of contracting the disease at any moment and 34.9% reported that sometimes they feel that they have been infected.

**Table 2:** Sources of distress

|  | Sources of stress |  |  |  |
| --- | --- | --- | --- | --- |
|  | Health of self, family and love-ones | Virus spread | Effects on daily life | Measures taken by the government |
| **Overall** | 0.07 ± 2.71 | 1.38 ± 1.90 | 2.37 ± 2.07 | 0.71 ± 1.95 |
| **Age** |  |  |  |  |
| 18-20 years | -0.19 ± 2.59** | 1.30 ± 1.93 | 2.32 ± 2.14 | 0.67 ± 1.89 |
| > 20-25 years | 0.14 ± 2.79 | 1.38 ± 1.87 | 2.36 ± 2.04 | 0.77 ± 1.97 |
| > 25-30 years | 0.96 ± 2.63 | 1.85 ± 1.65 | 2.48 ± 1.89 | 0.64 ± 1.98 |
| ≥ 31 years | 0.69 ± 2.47 | 1.37 ± 2.06 | 2.81 ± 1.75 | 0.68 ± 2.33 |
| **Gender** |  |  |  |  |
| Male | -0.47 ± 2.83 | 1.22 ± 2.00 | 2.28 ± 2.18 | 0.60 ± 2.10 |
| Female | 0.29 ± 2.62† | 1.44 ± 1.84 | 2.41 ± 2.01 | 0.76 ± 1.90 |
| **Province** |  |  |  |  |
| Punjab | 0.05 ± 2.70 | 1.36 ± 1.90 | 2.36 ± 2.07 | 0.72 ± 1.94 |
| Other provinces | 0.21 ± 2.85 | 1.60 ± 1.74 | 2.37 ± 1.93 | 0.73 ± 2.16 |
| **Education** |  |  |  |  |
| Medical | 0.33 ± 2.43 | 1.60 ± 1.98 | 2.32 ± 2.22 | -0.11 ± 2.32* |
| Pharmacy | 0.12 ± 2.71 | 1.44 ± 1.86 | 2.43 ± 1.99 | 0.76 ± 1.93 |
| Allied Health Sciences | -0.32 ± 2.78 | 1.22 ± 1.99 | 2.19 ± 2.31 | 0.90 ± 1.90 |
| Others | -0.02 ± 2.74 | 1.20 ± 1.94 | 2.21 ± 2.21 | 0.70 ± 1.95 |
| **Family member, relative or acquaintances got COVID-19** |  |  |  |  |
| Yes | 0.61 ± 2.66† | 1.66 ± 1.76** | 2.40 ± 2.10 | 0.83 ± 1.96 |
| No | -0.08 ± 2.70 | 1.30 ± 1.92 | 2.36 ± 2.05 | 0.68 ± 1.95 |

\*P-value < 0.05; \*\*P-value < 0.01; †P-value < 0.001; One-way ANOVA was applied for three or more groups. To estimate the difference among two groups independent sample t-test was applied. P-value < 0.05 was considered statistically significant.

### 3.4. Coping strategies

The overall brief-cope score of the respondents was 57.22 ± 12.29. As shown in Table 3, mean score was higher for religious coping (6.45 ± 1.68) followed by acceptance (5.58 ± 1.65) whereas it was the lowest for substance use (1.85 ± 1.35). Regarding the intra-demographic differences of coping strategies, there was no significant difference of the coping styles among age categories except for coping planning. Gender was observed to be one of the main factor where significant difference *(p* < 0.05) were observed for self-distraction, planning, acceptance and “religious coping”. Male respondents were observed to have a significantly lower score for the self-distraction, acceptance and religious coping while females had lower score for “planning” and humor coping. Amongst education categories, medical students had significantly less “self-blame” score than pharmacy *(p* = 0.017), allied health sciences *(p* = 0.011) and other university students *(p* = 0.008). Moreover, mean score of acceptance coping was significantly higher among those having family, relative, friend or acquaintance infected with COVID-19 than the ones who had not *(p* = 0.033).

**Table 3:** Coping Strategies adopted by the study participants

|  | Self-distraction | Active coping | Denial | Substance use | Emotional support | Informational support | Behavioral disengagement | Venting | Positive reframing | Planning | Humor | Acceptance | Religion | Self-blame |
| --- | --- | --- | --- | --- | --- | --- | --- | --- | --- | --- | --- | --- | --- | --- |
| **Overall** | 4.97 ± 1.61 | 4.81 ± 1.57 | 3.30 ± 1.59 | 1.85 ± 1.35 | 4.02 ± 1.67 | 3.62 ± 1.79 | 3.53 ± 1.54 | 3.79 ± 1.53 | 4.73 ± 1.71 | 4.63 ± 1.67 | 2.85 ± 1.40 | 5.58 ± 1.65 | 6.45 ± 1.68 | 3.11 ± 1.61 |
| **Age** |  |  |  |  |  |  |  |  |  |  |  |  |  |  |
| 18-20 years | 4.96 ± 1.59 | 4.66 ± 1.47 | 3.20 ± 1.59 | 1.77 ± 1.59 | 3.97 ± 1.62 | 3.60 ± 1.74 | 3.50 ± 1.51 | 3.74 ± 1.54 | 4.57 ± 1.62 | 4.43 ± 1.63* | 2.70 ± 1.34 | 5.50 ± 1.63 | 6.49 ± 1.65 | 3.07 ± 1.58 |
| > 20-25 years | 5.02 ± 1.63 | 4.89 ± 1.59 | 3.32 ± 1.59 | 1.94 ± 1.46 | 4.02 ± 1.68 | 3.62 ± 1.84 | 3.60 ± 1.58 | 3.83 ± 1.52 | 4.82 ± 1.76 | 4.69 ± 1.67 | 2.92 ± 1.46 | 5.60 ± 1.66 | 6.39 ± 1.72 | 3.22 ± 1.68 |
| > 25-30 years | 4.68 ± 1.64 | 5.11 ± 1.76 | 3.50 ± 1.56 | 1.58 ± 1.01 | 3.88 ± 1.59 | 3.56 ± 1.76 | 3.33 ± 1.51 | 3.86 ± 1.49 | 5.08 ± 1.86 | 5.09 ± 1.74 | 2.73 ± 1.23 | 5.89 ± 1.75 | 6.45 ± 1.66 | 2.70 ± 1.22 |
| ≥ 31 years | 4.53 ± 1.39 | 4.81 ± 1.67 | 3.75 ± 1.39 | 1.63 ± 1.15 | 4.78 ± 1.80 | 4.03 ± 1.76 | 3.22 ± 1.26 | 3.56 ± 1.26 | 4.91 ± 1.37 | 5.38 ± 1.64 | 3.00 ± 1.32 | 5.78 ± 1.66 | 6.94 ± 1.31 | 2.66 ± 1.09 |
| **Gender** |  |  |  |  |  |  |  |  |  |  |  |  |  |  |
| Male | 4.67 ± 1.57* | 4.70 ± 1.63 | 3.44 ± 1.58 | 1.95 ± 1.44 | 3.99 ± 1.62 | 3.65 ± 1.82 | 3.59 ± 1.56 | 3.73 ± 1.51 | 4.58 ± 1.75 | 4.79 ± 1.78 | 3.24 ± 1.60 | 5.39 ± 1.75* | 6.13 ± 1.84* | 3.19 ± 1.62 |
| Female | 5.09 ± 1.62 | 4.86 ± 1.54 | 3.24 ± 1.59 | 1.81 ± 1.31 | 4.03 ± 1.69 | 3.61 ± 1.78 | 3.51 ± 1.53 | 3.81 ± 1.53 | 4.80 ± 1.68 | 4.56 ± 1.62* | 2.68 ± 1.27* | 5.66 ± 1.61 | 6.58 ± 1.59 | 3.08 ± 1.61 |
| **Province** |  |  |  |  |  |  |  |  |  |  |  |  |  |  |
| Punjab | 4.96 ± 1.61 | 4.80 ± 1.55 | 3.28 ± 1.57 | 1.87 ± 1.35 | 4.02 ± 1.65 | 3.61 ± 1.77 | 3.54 ± 1.53 | 3.79 ± 1.51 | 4.73 ± 1.70 | 4.60 ± 1.66 | 2.83 ± 1.37 | 5.56 ± 1.65 | 6.43 ± 1.68 | 3.11 ± 1.59 |
| Other provinces | 5.05 ± 1.54 | 4.89 ± 1.75 | 3.49 ± 1.72 | 1.61 ± 1.26 | 4.01 ± 1.80 | 3.73 ± 1.98 | 3.43 ± 1.68 | 3.79 ± 1.65 | 4.81 ± 1.81 | 4.95 ± 1.81 | 3.16 ± 1.73 | 5.77 ± 1.74 | 6.73 ± 1.68 | 3.13 ± 1.83 |
| **Degree** |  |  |  |  |  |  |  |  |  |  |  |  |  |  |
| Medical | 4.77 ± 1.41 | 4.63 ± 1.55 | 2.84 ± 1.04 | 1.70 ± 1.41 | 3.91 ± 1.67 | 3.21 ± 1.84 | 3.07 ± 1.37 | 3.65 ± 1.60 | 4.33 ± 1.58 | 4.44 ± 1.45 | 3.14 ± 1.54 | 5.58 ± 1.68 | 6.09 ± 1.95 | 2.51 ± 1.03* |
| Pharmacy | 4.98 ± 1.61 | 4.82 ± 1.56 | 3.35 ± 1.59 | 1.91 ± 1.37 | 4.01 ± 1.65 | 3.68 ± 1.79 | 3.55 ± 1.56 | 3.81 ± 1.55 | 4.76 ± 1.71 | 4.65 ± 1.69 | 2.78 ± 1.34 | 5.56 ± 1.65 | 6.47 ± 1.68 | 3.10 ± 1.59 |
| Allied Health Sciences | 5.10 ± 1.68* | 4.85 ± 1.60 | 3.10 ± 1.50 | 1.76 ± 1.48 | 3.92 ± 1.43 | 3.66 ± 1.65 | 3.53 ± 1.47 | 3.79 ± 1.49 | 4.76 ± 1.59 | 4.71 ± 1.71 | 2.98 ± 1.53 | 5.35 ± 1.60 | 6.26 ± 1.74 | 3.29 ± 1.78 |
| Others | 4.94 ± 1.63 | 4.80 ± 1.57 | 3.26 ± 1.64 | 1.71 ± 1.22 | 4.08 ± 1.76 | 3.52 ± 1.82 | 3.54 ± 1.54 | 3.75 ± 1.44 | 4.71 ± 1.73 | 4.56 ± 1.64 | 2.97 ± 1.49 | 5.67 ± 1.66 | 6.47 ± 1.61 | 3.22 ± 1.68 |
| **Family member, relative or acquaintances got COVID-19** |  |  |  |  |  |  |  |  |  |  |  |  |  |  |
| Yes | 5.02 ± 1.65 | 4.83 ± 1.55 | 3.14 ± 1.48 | 1.99 ± 1.52 | 4.09 ± 1.74 | 3.69 ± 1.86 | 3.56 ± 1.57 | 3.82 ± 1.44 | 4.78 ± 1.76 | 4.64 ± 1.64 | 2.85 ± 1.38 | 5.78 ± 1.57 | 6.42 ± 1.64 | 3.22 ± 1.66 |
| No | 4.95 ± 1.60 | 4.81 ± 1.56 | 3.34 ± 1.61 | 1.81 ± 1.29 | 4.00 ± 1.64 | 3.60 ± 1.77 | 3.52 ± 1.53 | 3.78 ± 1.54 | 4.72 ± 1.69 | 4.62 ± 1.68 | 2.84 ± 1.40 | 5.52 ± 1.67* | 6.45 ± 1.69 | 3.08 ± 1.59 |

\*significant difference, One-way ANOVA was applied for three or more groups. To estimate the difference among two groups independent sample t-test was applied. P-value <0.05 was considered statistically significant.

## 4. Discussion

COVID-19 is the most devastating and challenging public health crises since the influenza pandemic in 1918. As of 14^th^ May 2020, more than 4.2 million people have been infected and 292046 succumbed to it globally (WHOa, 2020). It has brought pain and suffering to all the nations. It is undeniable that the disease is causing great deal of anxiety, fear and unrest in people of all ages. University students are no exception as it has been almost two months since the closure of all education institutions due to COVID-19 in Pakistan (Order No. SO(I&C-I) 1-2/2020). To the best of our knowledge, this is the first study that examine not only the psychological impact of the COVID-19 on university students but also their coping strategies. Regarding the anxiety and depression, a cut-point of ≥ 10 on GAD-7 as well as PHQ-9 is considered a “yellow flag” (drawing attention to a possible clinically-relevant condition), while a cut-point of 15 is a “red flag” (individuals in whom active treatment is probably warranted). In the present study, around 34% [GAD-7: moderate anxiety = 19.8%, severe anxiety = 13.7%) and 45% [PHQ-9: moderate = 21%, moderately severe = 13.6%, severe depression = 10.4%] respondents were found to have scores ≥ 10 on both aforementioned measures, respectively. Contrary to our results, Cao et al. (2020) reported that 21.3%, 2.7% and 0.9% of Chinese college students having mild, moderate and severe anxiety, respectively. Significantly higher proportion of students with anxiety and depression in our study can be attributable to the fact that 21.8% of our study subjects having somebody (family, friends, relatives, neighbors, acquaintance) who have been diagnosed with COVID-19 which was less than 1% in the previous study (Cao et al., 2020). Studies that assessed the psychological impact of SARS and MERS coronaviruses outbreaks also found significant impact of those epidemic on mental health of students (Wong et al., 2007; Al-Rabiaah et al., 2020). Upon enquiring the impact of anxiety and/or depressive symptoms on the quality of life, 48%, 11.6% and 6.5% stated somewhat, very and extreme difficulty in doing work, taking care of things at home, or getting along with others. It is need of the hour that academic institutions must work together with the government to promote measures suggested by the World Health Organization (2020) in order to improve the mental health of their students during CVOID-19. Following are the advices that can be useful in taking care of mental health during these unprecedented times (WHOb, 2020);

- Listen to advice and recommendations from the authorities.
- Have a healthy daily routine (wake up and go to sleep at similar times every day, keep up with personal hygiene, eat healthy, Exercise regularly, Allocate time for working and time for resting, and make time for doing things you enjoy)
- Try to minimize how much you watch, read or listen to news that makes you feel anxious or distressed. Seek the latest information at specific times of the day, once or twice a day if needed.
- As social contact is important to reduce stress, stay connected with people close to you by telephone and online channels.
- Avoid using alcohol and drugs as a way of dealing with fear, anxiety, boredom and frustration, and social isolation.
- Try to maintain a balance with on-screen and off-line activities in your daily routine.
- Use social media to promote positive and hopeful stories. Notably, correct misinformation related to COVID-19 wherever you see it.
- Offer support to others who may need it.

Eisenberg et al. (2012) reported two major components namely “avoidant coping” and “approach coping” in the Brief-COPE. As the humor and religion subscales did not exclusively load on either of the aforementioned factors, they were not included in either. Avoidant Coping is described by the brief-cope subscales of denial, substance use, venting, behavioral disengagement, self-distraction, and self-blame. It is not ideal at managing anxiety and has been linked with poorer physical health among those with medical conditions (Eisenberg et al., 2012). On the other hand, approach coping is characterized by the subscales of active coping, positive reframing, planning, acceptance, seeking emotional support, and seeking informational support. Compared to avoidant coping, it has been associated with better responses to adversity, including adaptive practical adjustment, better physical health outcomes and more stable emotional responding. However, Meyer (2001) categorized the strategies measured by the Brief-COPE into maladaptive coping and adaptive coping. Among other afore-mentioned subscales, religion as well humor were also considered as adaptive coping. Religious coping is defined as “religiously framed cognitive, emotional, or behavioral responses to stress, encompassing multiple methods and purposes as well as positive and negative dimensions” (Wortmann, 2016). In the present study, around 92% (doing this a lot = 56.9%, a moderate amount = 20.8%, a little bit = 14.2%) reported they have been trying to find comfort in their religion or spiritual beliefs and 93.3% (doing this a lot = 47.8%, a moderate amount = 29.5%, a little bit = 16%) reported that they have been praying or meditating. All in all, it was encouraging to see that scores for positive/adaptive coping strategies were greater than avoidant or maladaptive coping in our respondents.

This study had some limitations. Firstly, this study was conducted among the students of four higher education institutions. Secondly, as this was a web-based survey the problem of selective participation and coverage error might be present. Thirdly, we used a self-administered questionnaire so disadvantages associated with self-report data (introspective ability, response bias, sampling bias) could exist. Lastly, the clinical assessment for the diagnosis of depression and anxiety disorders as per criteria of Diagnostic and Statistical Manual of Mental Disorders (DSM-V) was not done. However, our findings provide valuable insight about the psychological impact of COVID-19, at its peak, on Pakistani university students.

## 5. Conclusions

COVID-19 pandemic have significant adverse impact on the mental health of Pakistani university students; prevalence of moderate to severe anxiety 34% and 24% students with moderately severe to severe depression. Major coping strategies adopted by the students are religious and acceptance coping. Our findings highlight that mental health should not be neglected during the epidemics. Educational institutions should work together with the authorities to promote measures suggested by the World Health Organization to improve mental health of their students.

## Data Availability

Data of the present study are available from the corresponding author on reasonable request

## Acknowledgement

Authors are grateful to the study participants for sparing their valuable time to fill-out the questionnaires.

## Authors’ contributions

MS, TMK, NS and KH conceived and designed the study. MS, NS, ZUM, MHR and MTK conducted literature review. MS, TMK, MHR and NA designed the questionnaire. MS, NS, HT and TMK analyzed and interpreted data. MS drafted the manuscript. TMK and KH, critically revised the manuscript. NS prepared the graphical abstract. All authors’ approved the manuscript for submission.

## Ethical considerations

This study was approved by the Research Ethics Committee of the Department of Pharmacy Practice, Faculty of Pharmacy, The University of Lahore, 1-Km Defense Road, Lahore, Pakistan.

